# Patterns of infection among travellers to Singapore arriving from mainland China

**DOI:** 10.1101/2023.01.16.23284584

**Authors:** Rachael Pung, Adam J. Kucharski, Zheng Jie Marc Ho, Vernon J. Lee

## Abstract

In light of the rapid growth of COVID-19 in mainland China, countries and regions outside of China have implemented travel restrictions of varying intensity. Using surveillance data of symptomatic travellers arriving from mainland China and detected in Singapore, this provides a proxy on the COVID-19 in mainland China. Furthermore, this allows us to ensure that travel-related restrictions commensurate with the current epidemiological situation and risk.

## Intro

During 2022, many COVID-19 control measures have been relaxed globally, including travel restrictions [1]. Global dynamics during 2020 were typically driven by local control efforts, which varied substantially, but this has gradually transitioned to dynamics driven by the emergence and spread of novel SARS-CoV-2 strains, combined with varying levels of population immunity. However, countries that have suppressed transmission for longer – such as mainland China – only encountered their first large nation-wide wave in late 2022. As a result, Omicron epidemic dynamics in these largely unexposed populations are likely to be considerably different to those underway elsewhere in the world.

This heterogeneity creates multiple challenges. First, there is a need to understand the local transmission dynamics in places that have relaxed public health measures and social interactions later than others, and how these outbreaks differ from other waves globally, particularly as local testing protocols change over time. In turn, there is a need to ensure that the global response to these outbreaks, including any travel-related interventions, are commensurate with the current epidemiological situation and risk. Throughout the COVID-19 pandemic, data on infections identified among travellers has provided crucial real-time situational awareness on international outbreaks [2, 3]. To inform planning and response to outbreaks in mainland China in 2022/23, we therefore used data on infections identified in Singapore among recent travellers from China, and estimated how infection dynamics in the country of origin changed over time.

## Methods

In Singapore, a confirmed case of COVID-19 is defined as an individual who tests positive via an antigen rapid test (ART) or PCR test administered by the healthcare provider. These cases are notified to the Ministry of Health and cases with a travel history in the 5 days prior to diagnosis are classified as imported cases. Since Apr 2022, travellers have not been required to take an on-arrival test for COVID-19 and only non-fully vaccinated travellers aged 13 and above are required to take a pre-departure test.

The incidence of imported travellers by the date of notification is not equivalent to the outbreak incidence in the country of origin because of the delay from infection to symptoms onset to testing to notification, and travellers demographics and risk of acquiring COVID-19 may not be similar to the local general population. We therefore adjusted the data to estimate the likely incidence of infection among travellers, and assumed that traveller infections reflect the shape of the epidemiological curve including its peak. For the Omicron variant, the mean incubation is 3.4 days (95% CI 2.88–3.96) [4]; in addition, there will be a brief delay between symptom onset and subsequent testing. We therefore assumed a mean delay of 5 days from infection to testing among imported cases and shifted the epidemic curve by this delay to estimate the expected number of infections per day [5]. Normalising by arriving traveller volume, we then estimated the incidence of COVID-19 per 1000 daily incoming travellers from mainland China and Hong Kong as a proxy for the outbreak situation in the respective regions. As a sensitivity analysis, we also generated the results using a delay of 4 days from infection to testing (i.e. no delay from symptoms onset to testing).

## Results

From 1 Nov 2022 to 5 Jan 2023, the average number of daily arrivals from mainland China was 553 (IQR 395–671). There were 207 imported cases in total identified from mainland China and all detected within 5 days of arrival. Two cases, aged 80 and above, from mainland China were hospitalised of which one was admitted into ICU. From 1 Dec 2022, from travellers’ data, we estimated that the outbreak in mainland China grew at a rate of 0.16 per day (i.e. doubling time of 4.3 days) and peaked around 15 Dec 2022 (Fig 1A). From 15 Nov 2022 to the peak of the outbreak, we estimated a cumulative attack rate of 14% among travellers, and by the end of 2022, it was 31% (Fig 1B).

**Figure 1:**
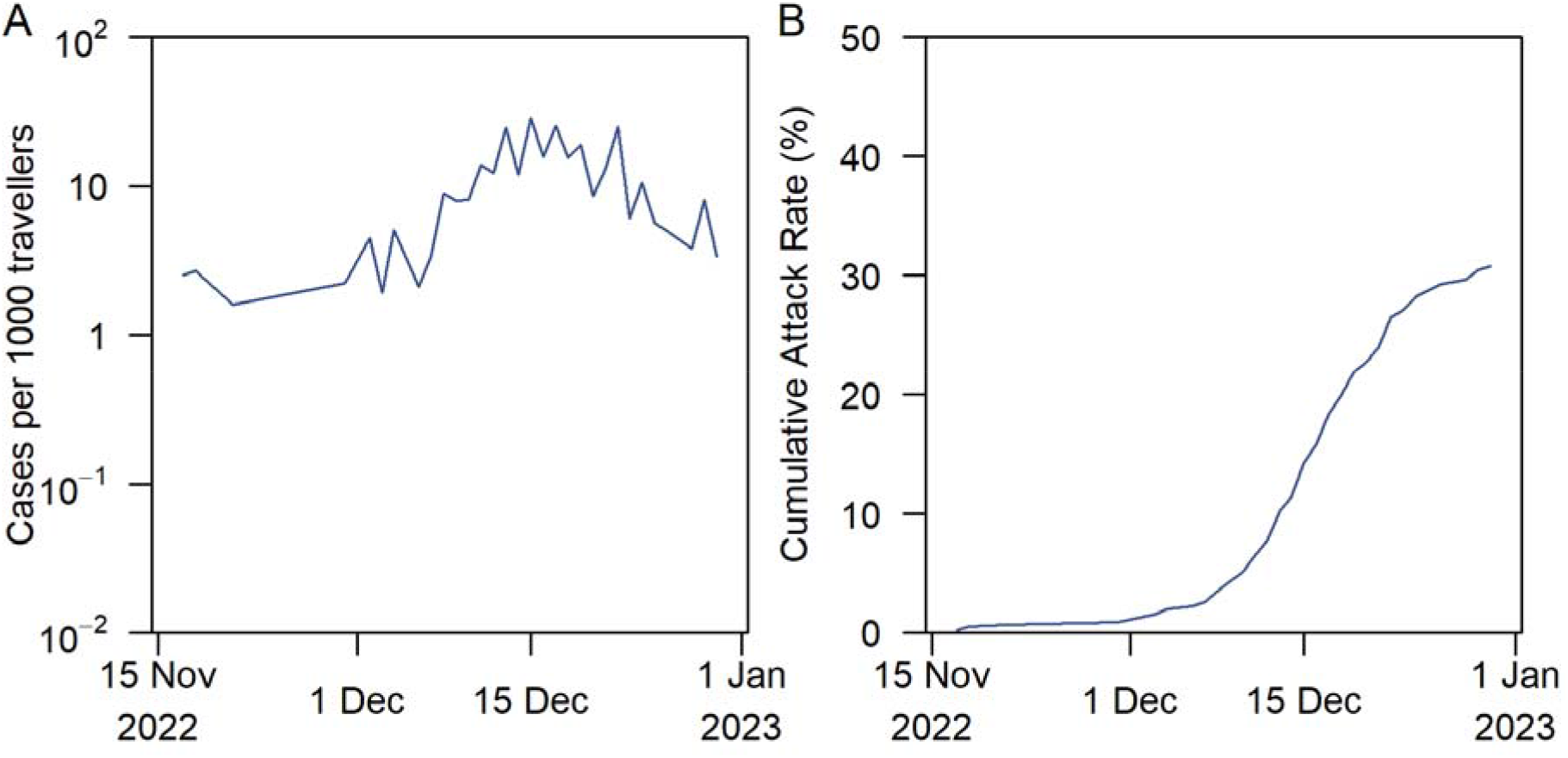
COVID-19 outbreak metrics for mainland China. (A) Estimated number of COVID-19 cases per 1000 travellers arriving from mainland China and (B) cumulative attack rate.

As a sensitivity analysis, we omitted 3 imported cases from mainland China as these cases had a positive test after 5 or more days since their date of arrival, given they may have been infected outside their country of origin. Using this subset of data, we estimated a lower outbreak growth rate of 0.15 per day (i.e. doubling time of 4.5 days) in mainland China, with the peak occurring on 16 Dec 2022, and a lower cumulative attack rate of 11% at the height of the outbreak (Supplementary Fig 1).

## Discussion

Using locally reported cases in Singapore with a known travel history to mainland China, we estimated the number of arriving infections over time from these two regions. Our results show the benefits of combining epidemiological information, including date of notification and symptoms status, with travel movements to understand importations as well as dynamics internationally.

We estimated that infection incidence among travellers from mainland China peaked at around 29 cases per 1000 travellers per day (i.e. 2.9%) in mid-December 2022. For comparison, estimates based on community testing in the UK suggested a peak in incidence of just under 1% in early 2022 [6]. This is consistent with the larger initial reproduction number observed in mainland China, which all things being equal would typically lead to a shorter epidemic with a larger peak.

There are some limitations to our analysis. Local testing approaches may potentially miss asymptomatic cases, which would mean cumulative attack rate among travellers would be higher in reality. Our estimate of 30% attack rate up to the end of 2022 is lower than the estimated 75% attack rate estimated for Beijing [7]. However, this difference may also be down to regional differences in mainland China and/or travellers being non-representative of the wider population. There could also be heterogeneity in other demographics, for example models using traveller incidence which reflect the proportion of travellers from urban cities compared to rural areas. The data we analyse from Singapore also contains no information on patterns of under-ascertainment over time (e.g. as measured by contact tracing and proactive case finding) but if the proportion of infections is reasonably constant during the period analysed, under-ascertainment would not affect the estimates of the growth rate and time of peak. Testing behaviour among travellers may also have changed over time, but this is unlikely to be a major issue for Singapore since no travel measures were introduced or removed during the period of interest.

Despite the widespread deployment of travel restrictions by many countries, such as pre-departure and post-arrival testing, in December 2022 in response to the wave in mainland China, the impact on transmission in those countries is likely to be limited. During a growing epidemic, most infections would have been recent, which means that many travellers will be incubating or in the early stage of infection when they are less likely to test positive. Moreover, we estimated that incidence among travellers from mainland China were already peaking by the time many countries introduced measures in mid-December 2022. The level of traveller incidence also suggests that the absolute number of infected travellers would have been many times smaller than the number of locally reported cases during the same period in many countries introducing such measures. This is especially since flight volumes out of China were low over this period.

As well as missing infections among travellers, testing at point of departure does not provide situational awareness for the country of arrival. In contrast, general testing of incoming travellers, either at the border for those arriving from countries with ongoing outbreaks [8] or following subsequent symptom onset, as in the analysis prevented here, can provide insights into the situation from across the world. As a long-term measure, there may be challenges with estimating importation patterns from locally detected cases if testing behaviours among travellers were to change. Thus, surveillance and control efforts in a country should be tailored to the epidemiological situation and current COVID-19 response objectives in a given country [9], and traveller surveillance will be a good adjunct tool.

## Data Availability

All data produced in the present work are contained in the manuscript

## Funding Additional Information

RP acknowledges funding from the Singapore Ministry of Health. AJK was supported by a Sir Henry Dale Fellowship jointly funded by the Wellcome Trust and the Royal Society (grant 206250/Z/17/Z). The funders had no role in study design, data collection and interpretation, or the decision to submit the work for publication.

## Supplementary Info

**Supplementary Figure 1:**
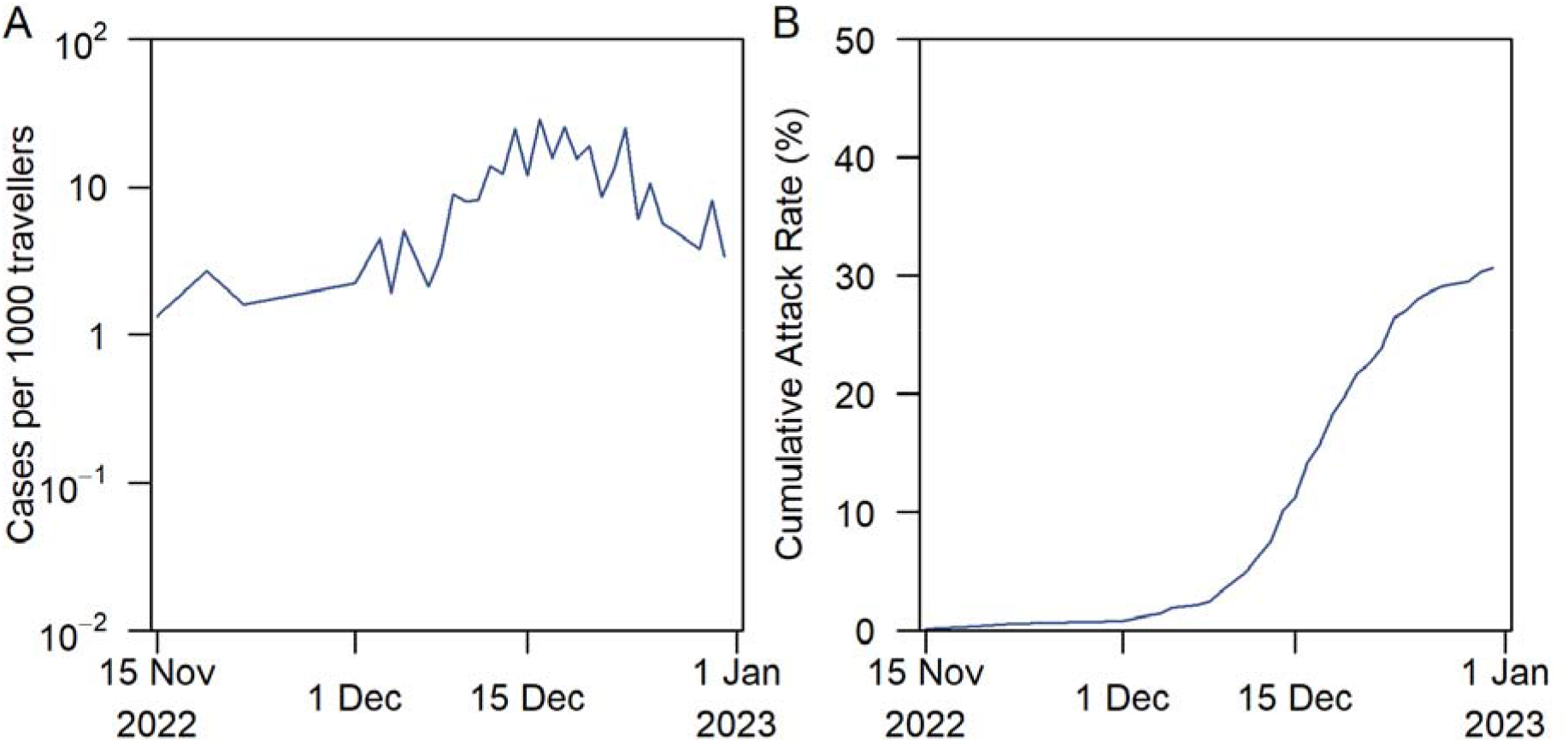
COVID-19 outbreak metrics for mainland China. (A) Estimated number of COVID-19 cases per 1000 travellers arriving from mainland China and (B) cumulative attack rate,

